# How long is the long COVID? a retrospective analysis of football players in two major European Championships

**DOI:** 10.1101/2023.03.11.23287138

**Authors:** Sandra Miccinilli, Marco Bravi, Giorgio Conti, Federica Bressi, Silvia Sterzi, Fabio Santacaterina, Massimo Ciccozzi

**Affiliations:** Department of Physical and Rehabilitation Medicine, Università Campus Bio-Medico di Roma, Via Alvaro del Portillo, 200, 00128 Rome, Italy; Unit of Medical Statistics and Molecular Epidemiology, Università Campus Bio-Medico di Roma, Via Alvaro del Portillo, 200, 00128 Rome, Italy

**Author notes:** the authors Marco Bravi and Sandra Miccinilli contributed equally to this paper.

**Keywords:** SARS-CoV-2, muscle injury, soccer, muscle strain

## Abstract

**Objectives:** the goal of this study was to investigate the correlation between SARS-CoV-2 infection and muscle injuries among a large sample of professional soccer players.

**Methods:** A retrospective cohort study was conducted on professional soccer players from the Serie A and LaLiga leagues during the 2019-2020 and 2020-2021 football seasons. The players were divided into two groups based on whether they contracted the Sars-CoV-2 infection (C+) or not (C-) during the 2020/2021 season. Data collection was conducted using the Transfermarkt24 site.

**Results:** In the 2019-2020 both championships showed non-significant differences in the average number of muscular injuries between the C+ group and the C- group (Serie A: p=0.194; 95%CI: - 0.044 to 0.215, LaLiga p=0.915; 95%CI: -0.123 to 0.137). In the 2020-2021 the C+ group had a significantly higher number of muscular injuries compared to the C- group in both championships (Serie A: p<0.001; 95%CI 0.731 to 1.038; LaLiga: p<0.001; 95%CI: 0.773 to 1.054). Multiple linear regression analysis confirmed that belonging to C+ in the season 2020/2021 was the variable that most strongly influenced the probability of having a muscle injury in both championships. Survival analysis revealed a hazard ratio of 3.73 (95%CI 3.018 to 4.628) and of 5.14 (95% CI 3.200 to 8.254) for Serie A and LaLiga respectively.

**Conclusions:** This retrospective cohort study revealed a significant association between SARS-CoV-2 infection and increased risk of muscle injury, emphasizing the importance of carefully considering the infection in the decision-making process for determining athletes’ readiness to return to sport.

## Introduction

In late December 2019, The World Health Organization (WHO) office in China was informed about novel cases of pneumonia of unknown aetiology detected in the city of Wuhan, Hubei province [1]. Afterwards, a new type of coronavirus, named SARS-CoV-2, was isolated and identified by the Chinese authorities. The coronavirus disease 2019 (COVID-19) caused by SARS-CoV- 2 was classified as a pandemic on 11 of March 2020 [2] by the World Health Organization (WHO). All countries were involved in surveillance of the disease and all sports that included close contact were temporarily suspended.

Football (soccer) is a physically demanding sport that requires a high level of neuromuscular readiness, including strength, reactivity, and muscular power [3,4]. The characteristics of this sport could therefore justify the increases in muscle injury rates associated with the increase in the number of official seasonal competitions and weekly training sessions [5,6]. However, the outbreak of the SARS-CoV-2 virus has had a significant impact on professional athletes, even in mild cases, leading to muscle weakness, poor tolerance to physical exercise, period of inactivity and absence from sporting practice, with a considerable impact also on bio-psychosocial context [7–9]. COVID-19 is known to cause severe inflammatory responses, respiratory failure, acute respiratory distress syndrome (ARDS), and bilateral pneumonia [10]; in some cases these symptoms can continue causing what is called long COVID. It can also affect the musculoskeletal system, causing myalgia and sarcopenia in COVID-19 positive patients [11]. Therefore, the question arises whether COVID- 19 may have influenced the number of muscle injuries in elite athletes. Previous studies in the literature have observed that the approximately three months of sports activity suspension due to the lockdown caused an increase in the incidence of injuries in the main European football leagues, including the Bundesliga (GER) [12], LaLiga (SPA) [13] and Premier League (ENG) [14]. The only exception is represented by Italian first division (Serie A) [15], where a non-statistically significant difference was observed between pre- and post-lockdown injuries. However, these studies did not show whether contracting SARS-CoV-2 infection can be considered an additional risk factor for injury occurrence. Only the recent prospective study by Wezenbeek et al. [16] involving three Belgian professional male football teams during the first half of the 2020-2021 season, reported a five-fold higher risk of developing a muscle strain after a SARS-CoV-2 infection. The goal of this study is to further investigate the findings reported by Wezenbeek et al. [16] and to retrospectively examine, on a larger sample size, the correlation between SARS-CoV-2 infection and muscle injuries among professional football players from two different championships, Serie A and LaLiga. These two championships were chosen because Serie A was an exception in terms of post-lockdown injury increase, while LaLiga had the most active collaboration in identifying and communicating positive cases and its results were more discordant between pre- and post-lockdown.

However, these studies did not show whether having contracted SARS-CoV-2 infection can be considered an additional risk factor for occurrence of injuries. Recently only the prospective study by Wezenbeek et al., involving three Belgian professional male football teams, during the first half of the 2020–2021 season reported a five times higher risk of developing a muscle strain after a SARS-CoV-2 infection.

The goal of this study is to deepen the results reported by Wezenbeek et al. and to retrospectively verify, on a larger sample, the correlation between SARS-CoV-2 infection and muscle injuries among professional footballers of two different championship Serie A and LaLiga. The choice of these two different championships was because Serie A represented the exception of post-lockdown injuries increase and both Serie A and LaLiga had the most active collaboration in identifying and communicating positive cases [13]. Our hypothesis is that there is a relationship between muscle injury and previous SARS-CoV-2 infection.

## Materials and methods

### Study design

A retrospective cohort study was conducted according to the STROBE guidelines [17]. The aim was to identify a potential correlation between SARS-CoV-2 infection and muscle injuries in professional soccer players from the Serie A and LaLiga leagues during the 2019-2020 and 2020-2021 football seasons. The study included all players from Serie A and LaLiga during the two seasons, as well as those who changed league levels during this time period (e.g. from a minor league such as Serie B in Italy to the top national league, Serie A) and those who moved to a foreign team. Athletes were divided into two groups: COVID-positive (C+) and COVID-negative (C-).

The grouping was performed as follows: 1) all football players infected during the 2020-2021 season were classified as “C+”, while all players who did not contract the infection during this time period were classified as “C-”; 2) players who contracted the infection during the 2019-2020 season or during off-season periods (e.g. season summer suspension) were excluded from the study (Serie A n = 29; LaLiga n = 38) and their data were removed from both seasons. This created a database with pre-COVID (2019-2020 season) and post-COVID (2020-2021 season) data.

### Patient and public involvement

This study investigating the association between SARS-CoV-2 infection and the occurrence of muscle injuries during the sports season is the result of the involvement of medical staff and young athletes from some sports clubs who were the first to notice this possible correlation. These have also been involved in the discussion of the results of the study.

### Data collection

Data collection was conducted using the Transfermarkt24 site (https://www.transfermarkt.co.uk/) a website founded in Germany in May 2000 which contains a wealth of football information from leagues around the world, including data on the type and severity of injuries. This methodology is consistent with previous studies [18–21]. The data collected are specified in Table 1.

**Table 1:** data collected and relative definitions used in the study

|  |  |
| --- | --- |
| Birth date |  |
| Age at the end of the 2020-2021 season |  |
| Team |  |
| Muscle injuries season 2019-2020 | Total number of muscular injury sustained by a player that resulted from a in-season football match or football training |
| Muscle injuries season 2020-2021 |  |
| Matches played in the 2019-2020 | Total number of matches played during the season |
| Matches played in the 2020-2021 |  |
| Minutes played season 2019-2020 | Total number of minutes played during the season |
| Minutes played season 2020-2021 |  |
| Date of SARS-CoV-2 infection | Date of in-season SARS-CoV-2 infection |
| Date of muscle injuries season 2019-2020 | Date of each muscle injury occurred during in-season period |
| Date of muscle injuries season 2020-2021 |  |
| Season 2019-2020 start and end date | First and last match date of the season |
| Season 2020-2021 start and end date |  |

### Statistical analysis

Statistical analysis was performed according to the CHAMP statement [22] using MedCalc software (Version 20, MedCalc Software Ltd). The normal distribution of the data was verified using the Kolmogorov-Smirnov test. The comparison of mean injuries between the two groups in the 2019- 2020 and 2020-2021 seasons was performed using a parametric t-Student test. A multiple linear regression analysis was carried out in both seasons to analyse the relationship between the dependent variable, muscle injuries, and the independent variables of age, SARS-CoV-2 infection, minutes played, and matches played. A multiple linear regression analysis was carried out in both seasons to analyse the relationship between the dependent variable, muscle injuries, and the independent variables of age, SARS-CoV-2 infection, minutes played, and matches played. Additionally, a Kaplan-Meier survival analysis was performed, using the elapsed time (in days) from the start of each championship to the day on which the injury occurred during the in-season period as the “survival time.” If no injuries occurred, the total duration of the season in days was used as the survival time. The Kaplan-Meier curves were analysed using the Logrank Test in both seasons.

## Results

The study included 634 players from Serie A and 649 players from LaLiga, for a total of 1283 elite football players. Of the Italian championship players, 171 (27.3%) were infected with SARS-CoV-2, with 29 excluded due to being infected during the 2019-2020 season. In LaLiga, a total of 165 (25.4%) players were infected with SARS-CoV-2, with 38 excluded as they were infected during the 2019-2020 season (Figure 1).

**Figure 1:**
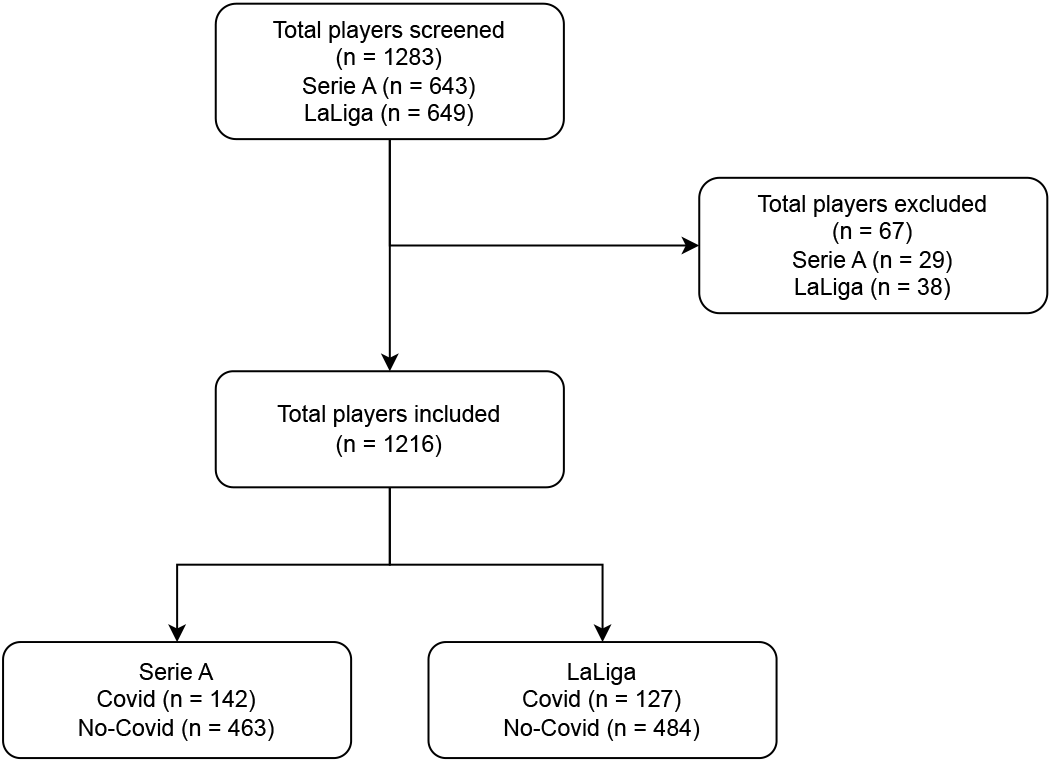
Participants enrolment flowchart

In the 2019-2020 Serie A championship, non-significant differences (p=0.194; 95%CI: -0.044 to 0.215) were found in the average number of muscular injuries between the C+ group (0.52±0.75) and the C- group (0.43±0.68). In the 2020-2021 season, the C+ group had a significantly higher number of muscular injuries (1.44±0.96) compared to the C- group (0.56±0.78) (p<0.001; 95%CI 0.731 to 1.038) (figure 2).The multiple linear regression analysis revealed that, in the 2019-2020 season, muscular injuries were mainly related to age, minutes played, and matches played by the athletes (table 2). In the 2020-2021 season, in addition to age, belonging to the C+ group was found to be the variable that most strongly influenced the probability of having a muscle injury, unlike the previous season (table 2).

**Table 2:** Multiple linear regression analysis of Serie A football players. The factors described, relating to the 2019-2020 and 2020/2021 season, represent the dependent variables capable of influencing or not the number of muscle injuries within the same season.

| Independent variables | Coefficient | Std. Error | t | P |
| --- | --- | --- | --- | --- |
| **2019/2020 season** |  |  |  |  |
| (Constant) | -0,123 |  |  |  |
| Age | 0,023 | 0,006 | 3,900 | **<0,001** |
| Minutes played | -0,000 | 0,000 | -3,679 | **<0,001** |
| Matches played | 0,025 | 0,007 | 3,613 | **<0,001** |
| Group (C+/C-) | -0,108 | 0,065 | -1,658 | 0,098 |
| **2020/2021 season** |  |  |  |  |
| (Constant) | 1,770 |  |  |  |
| Age | 0,020 | 0,007 | 2,814 | **0,005** |
| Minutes played | -0,000 | 0,000 | -1,269 | 0,205 |
| Matches played | 0,006 | 0,007 | 0,910 | 0,363 |
| Group (C+/C-) | -0,889 | 0,078 | -11,371 | **<0,001** |

**Figure 2:**
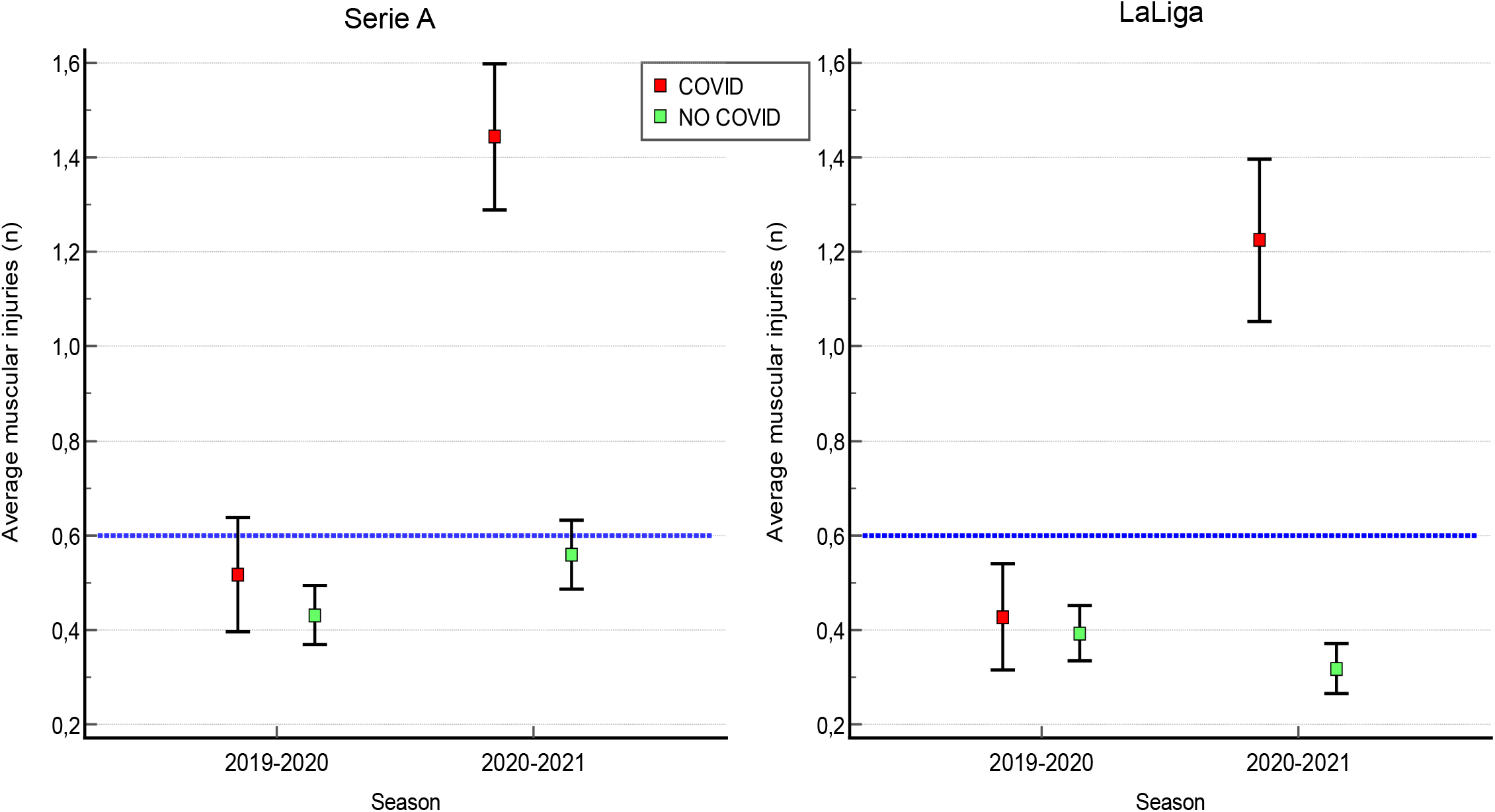
Average number of muscle injuries in the 2019/2020 and 2020/2021 seasons. The dashed blue line represents the average number of injuries per season identified by Bengtsson et al [25].

The survival analysis based on the Kaplan-Meier curves (figure 3) revealed that in both groups, the injury rates during the 2019-2020 season did not differ significantly (p=0.401). However, during the 2020-2021 season, the difference in injury rates between the two groups became statistically significant (p<0.001), with a hazard ratio of 3.73 (95%CI 3.018 to 4.628).

**Figure 3:**
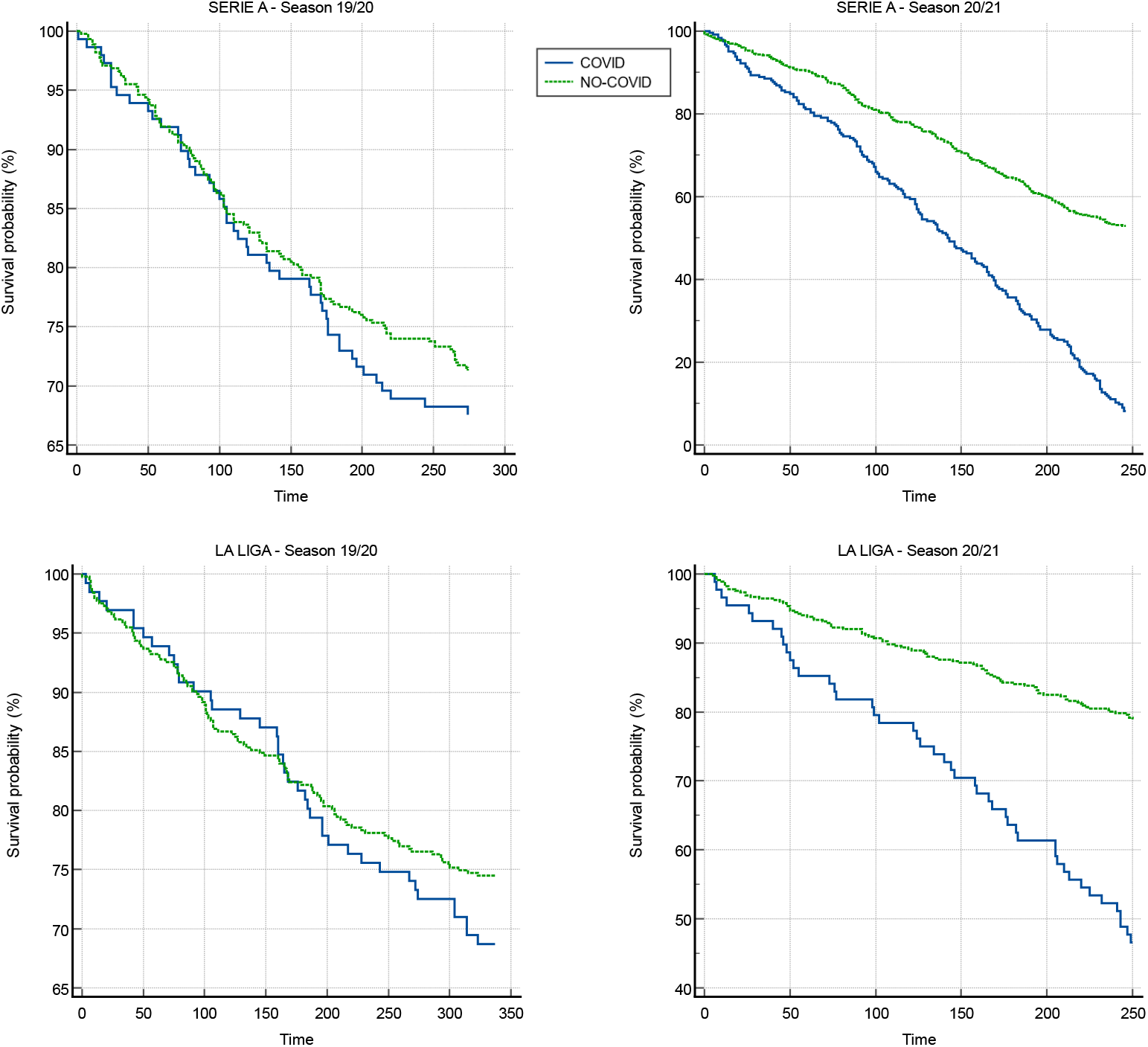
Kaplan-Meier curves relating to muscular injuries events of Serie A and LaLiga in the 2019-2020 and 2020-2021 seasons.

In LaLiga, the results indicated that during the 2019-2020 season, there was no significant difference in the average number of muscular injuries between the C+ and C- groups, with 0.40±0.67 and 0.39±0.65 injuries, respectively (p=0.915; 95%CI: -0.123 to 0.137). However, during the 2020-2021 season, the average number of injuries was significantly higher (p<0.001; 95%CI: 0.773 to 1.054) in the C+ group (1.23±1.04) compared to the C- group (0.32±0.59) (figure 2). The multiple linear regression analysis indicated that in the 2019-2020 season, age was the main factor related to muscle injuries (p=0.044), while games played and minutes played by athletes did not significantly influence the occurrence of injuries. However, in the 2020-2021 season, the group variable had a significant influence on the muscle injury rate (table 3).

**Table 3:** Multiple linear regression analysis of LaLiga football players. The factors described, relating to the 2019-2020 and 2020/2021 season, represent the dependent variables capable of influencing or not the number of muscle injuries within the same season.

| Independent variables | Coefficient | Std. Error | t | P |
| --- | --- | --- | --- | --- |
| **2019/2020 season** |  |  |  |  |
| (Constant) | 0,110 |  |  |  |
| Age | 0,0129 | 0,006411 | 2,022 | **0,044** |
| Minutes played | -0,000 | 0,00006924 | -0,426 | 0,670 |
| Matches played | 0,002 | 0,006129 | 0,337 | 0,736 |
| Group (C+/C-) | -0,042 | 0,06376 | -0,663 | 0,508 |
| **2020/2021 season** |  |  |  |  |
| (Constant) | 2,141 |  |  |  |
| Age | 0,000 | 0,007 | 0,087 | 0,931 |
| Minutes played | 0,000 | 0,000 | 0,697 | 0,486 |
| Matches played | -0,004 | 0,006 | -0,749 | 0,454 |
| Group (C+/C-) | -0,902 | 0,069 | -12,916 | **<0,001** |

Also in the Spanish championship, the survival analysis (figure 2) showed how the injuries rates in the 2019-2020 season did not differ significantly (p=0.246), while in the season 2020-2021 the difference becomes statistically significant (p< 0.001), with a hazard ratio of 5.14 (95% CI 3.200 to 8.254).

## Discussion

The main aim of this study was to determine whether professional soccer players who contracted SARS-CoV-2 had an increased risk of muscular injuries. The findings of this study reported a three to five times increased risk of muscle injury after SARS-CoV-2 infection.

Over the years, UEFA has conducted numerous studies on the match/minute ratio played by the elite athletes of the teams participating in its international tournaments to define a “normal” value for the incidence of injuries in football [23–25]. In particular, Hägglund et al.’s 11-year retrospective study [23] observed the incidence of injuries in more than 1000 hours of play and found an incidence of 2 injuries per year per individual footballer or an average of 50 injuries if we take into consideration the team group (list of 25 athletes). If we consider only muscular injuries, the incidence drops to 15 injuries out of a shortlist of 25 footballers, with an average per athlete of 0.6 injuries [25]. This data is essential for commenting on the results of our study. Our results showed that in both leagues, players who did not contract COVID-19 had injury rates lower than the average of 0.6 reported by Bengtsson et al. [25] in both seasons. Instead, regarding players infected with SARS-CoV-2, they showed in the pre-covid season an injury rate in line with the data of Bengtsson et al. [25], while in the 2020/2021 season, after the infection, it increased significantly in both leagues (1.44 for Serie A and 1.23 for LaLiga).

Our results indicate that SARS-CoV-2 infection could be a risk factor for muscular injury, in line with the recent study by Wezenbeek et al. [16]. Multiple linear regression analysis showed that in the 2019-2020 season, the risk of injury was significantly influenced by age, according to a previous study by Green et al. [26]. However, in the 2020-2021 season, SARS-CoV-2 infection was the variable that highly influenced the risk of muscle injury. Therefore, belonging to the C+ group in the 2020-2021 season completely distorts the correlation between the various variables and the muscle injury rates, becoming the main factor capable of influencing the latter. In other words, while older players with a high number of games and playing time were at greater risk of incurring muscle injuries in the 2019-2020 season, having contracted COVID-19 was the primary and most important risk factor in the following season. Similarly, in LaLiga, age was the main risk factor for muscle injury in the 2019-2020 season, while in the 2020-2021 season, the only variable that significantly influenced muscle injuries was the SARS-CoV-2 infection. The survival analysis showed that during the 2019- 2020 season, the two groups had a similar risk of muscular injury. In the 2020/2021 season, the hazard risk ratio was 3.7 to 5.1 times greater among those athletes infected by SARS-CoV-2, in line with the results of Wezenbeek et al. [16], which reported a five-time higher hazard rate to develop a muscle strain after SARS-CoV-2 infection.

As discussed by Wezenbeek et al. [16], there are two possible explanations for the correlation between SARS-CoV-2 infection and an increased risk of muscular injury. The first hypothesis is that strict quarantine rules implemented during the 2020/2021 season led to prolonged periods of abstention from training and sports participation, resulting in muscular detraining [27,28] and subsequent loss of muscle strength [29]. Muscle strength has been shown to be a protective factor for muscular injury [30], and the loss of muscle strength is more significant in highly trained athletes with greater initial muscle mass [31]. The second explanation is related to the direct biological effects of the virus. SARS-CoV-2 infection causes hyperinflammation, an increase in inflammatory markers, an increase in the neutrophil/lymphocyte ratio, and possible depletion of circulating T cells [32]. Furthermore, the virus may cause direct damage to muscle tissue by targeting the ACE 2 receptor [18], which is widely present on muscle tissue [33]. In addition, the virus can cause disturbances in blood flow and oxygen transport, leading to reduced muscle oxygenation during exercise. Lower muscle oxygen saturation [34] and VO2 max [35] are possible risk factors for muscle injury.

### Clinical Implications

These results emphasize the importance of carefully considering the infection in the decision-making process for determining athletes’ readiness to return to sport (RTS). In fact, as already described by Elliot et al. [36] in addition to the known complications, it is necessary to consider that COVID-19 can be responsible for musculoskeletal complications. Therefore, our opinion is that the decision regarding the RTS should not only take into account the remission of COVID-19 related symptoms, rest and the cessation of post-COVID-19 drug therapy, but should include specific assessment and training programs, generally used after a musculoskeletal injury [37] and therefore consider COVID-19 not only a risk factor but a real injury.

### Limitations

One possible limitation of this study is that the muscle injury events were collected from a single database, which may have resulted in some injuries being overlooked, omitted, or interpreted differently, leading to over- or underestimation. Another limitation of this study is that it was not possible to control for some factors that may have influenced the increased risk of injury, such as previous muscle injuries in unexamined seasons and severity of COVID-19-related symptoms. Nevertheless, the study’s strength lies in its design, with a large homogeneous population group of male professional soccer players followed prospectively with a standardized methodology.

## Conclusion

This retrospective cohort study, conducted on a large sample of professional male football players from two main European leagues, has revealed a significant association between SARS-CoV-2 infection and increased risk of muscular injury. The results of this study show that the risk of muscular injury is 3.7 to 5.1 times higher in football players who have been infected with SARS-CoV-2. Overall, our study highlights the need for further investigation into the effects of SARS-CoV-2 on athletes and the development of tailored rehabilitation protocols to ensure safe return to play.

## Data Availability

All data produced in the present study are available upon reasonable request to the authors

## Patient and public involvement

patients and/or the public were involved in this research. Refer to the Methods section for further details.

## Contributorship

Contributors MB, GC, and MC contributed to the conception of the study. SM, MB, GC, FB, SS, FS and MC contributed to the design of the study. GC, MB and FS completed the acquisition of the data. MB GC and MC performed the data analysis. All authors assisted with the interpretation. MB and SM were the principal authors of the manuscript. All authors contributed to the drafting and revision of the final article. All authors approved the final submitted version of the manuscript.

## Ethical approval information

Not applicable

## Acknowledgements

Not applicable

## Data availability

Data are available on reasonable request to the corresponding author.

